# iGTP: Learning interpretable cellular embedding for inferring biological mechanisms underlying single-cell transcriptomics

**DOI:** 10.1101/2024.03.29.24305092

**Authors:** Kang-Lin Hsieh, Kai Zhang, Yan Chu, Lishan Yu, Xiaoyang Li, Nuo Hu, Isha Kawosa, Patrick G. Pilié, Pratip K. Bhattacharya, Degui Zhi, Xiaoqian Jiang, Zhongming Zhao, Yulin Dai

## Abstract

Deep-learning models like Variational AutoEncoder have enabled low dimensional cellular embedding representation for large-scale single-cell transcriptomes and shown great flexibility in downstream tasks. However, biologically meaningful latent space is usually missing if no specific structure is designed. Here, we engineered a novel interpretable generative transcriptional program (iGTP) framework that could model the importance of transcriptional program (TP) space and protein-protein interactions (PPI) between different biological states. We demonstrated the performance of iGTP in a diverse biological context using gene ontology, canonical pathway, and different PPI curation. iGTP not only elucidated the ground truth of cellular responses but also surpassed other deep learning models and traditional bioinformatics methods in functional enrichment tasks. By integrating the latent layer with a graph neural network framework, iGTP could effectively infer cellular responses to perturbations. Lastly, we applied iGTP TP embeddings with a latent diffusion model to accurately generate cell embeddings for specific cell types and states. We anticipate that iGTP will offer insights at both PPI and TP levels and holds promise for predicting responses to novel perturbations.

## Introduction

With the rapid advent of single-cell technology, millions of cellular transcriptomes have been generated by large consortia [e.g. Human Cell Atlas^1^, Tabula Sapiens^2^] and various individual studies to advance our understanding of the human body, developmental biology, tumor heterogeneity, and immune system dynamics, leading to more precise diagnostic and therapeutic strategies^3^. The massive increase in cellular transcriptomes also provides opportunities to use deep-learning frameworks to learn and represent heterogeneous biological functions.

The deep-learning framework Variational AutoEncoder (VAE)^4^, has been widely adapted for dimension reduction^5^ and a variety of other applications^6,7,8^ in high-dimensional single-cell RNA-seq data analysis. By leveraging its probabilistic nature, VAE effectively addresses challenges such as batch effect correction^7^, imputation^9^, and perturbation response prediction^8,10^, which shows significant advantages over traditional statistical methods in terms of scalability and the ability to detect nonlinear patterns^11,12^. Deep generative models excel in their specific modeling tasks but often lack interpretability and struggle to provide biologically relevant latent representations of protein-protein interactions (PPI) and biological pathways. Without implementing specific model structures, these models cannot directly capture variations in biological processes^8^. Elmarakeby et al. (P-NET)^13^ introduced a new fashion of using fully connected layers to encode various levels of biological information, including genes, pathways, and biological processes, for outcome prediction, providing a biologically meaningful interpretation via the weight between layers. However, P-NET is primarily designed for prediction purposes and is not a generative model. More recently, methods such as single-cell embedded topic model (scETM)^14^ engineered the Decoder layer to ensure the embedding layers accurately represent gene and topic embeddings, as well as prior gene modules.

However, the topics from the scETM cannot be directly linked to biological meaning. The VAE Enhanced by Gene Annotations (VEGA)^15^ designed a VAE model to learn gene module variables (GMV) space via employing a sparse linear Decoder with masking to preserve a connection from a GMV to an output gene exclusively if the gene is recognized as part of the specific gene module. However, this one-layer Decoder simplified the linkage between genes and GMV, neglecting to incorporate prior knowledge like PPIs.

To advance beyond these constraints, we propose iGTP (interpretable generative transcriptional program), a VAE with a masked multi-layer linear Decoder informed by biological pathways/transcriptional programs (TPs) and protein-protein interactions. iGTP offers the following functions: 1) direct interpretable latent space for TPs related to biological state alteration and use these high dimensions to represent cellular embeddings; 2) uncover previously unknown associations between PPIs/TPs and cellular state alteration; 3) infer TP activity in unseen data using pretrained model; 4) infer gene expression alteration for unseen perturbation; 5) assess the relative importance for TPs and PPIs, providing new insights for the underlying biological mechanism; 6) demonstrate the generative capability of TP embeddings in creating new cells corresponding to specified cell types and condition.

## Results

### iGTP framework

We developed a customized VAE model to capture the biologically meaningful embedding in latent spaces for scRNA-seq data, enabling the interpretation of transcriptional shifts and leveraging Graph Neural Networks (GNN) for propagating gene alteration inferences through the PPI network (Fig. 1). To ensure a biologically interpretable latent layer, we engineered a sparse multilayer Decoder with prior biological knowledge of transcription programs and PPIs, which explicitly guides the model to learn the differential TP and PPI responsible for cellular states shift. Similar to the previous graph-enhanced gene activation and repression simulator (GEARS) model^16^, we further incorporate the latent TP embeddings with gene-perturbation PPI embedding to predict the perturbation response of TPs and genes on unseen scenarios of certain genes.

**Fig. 1.**
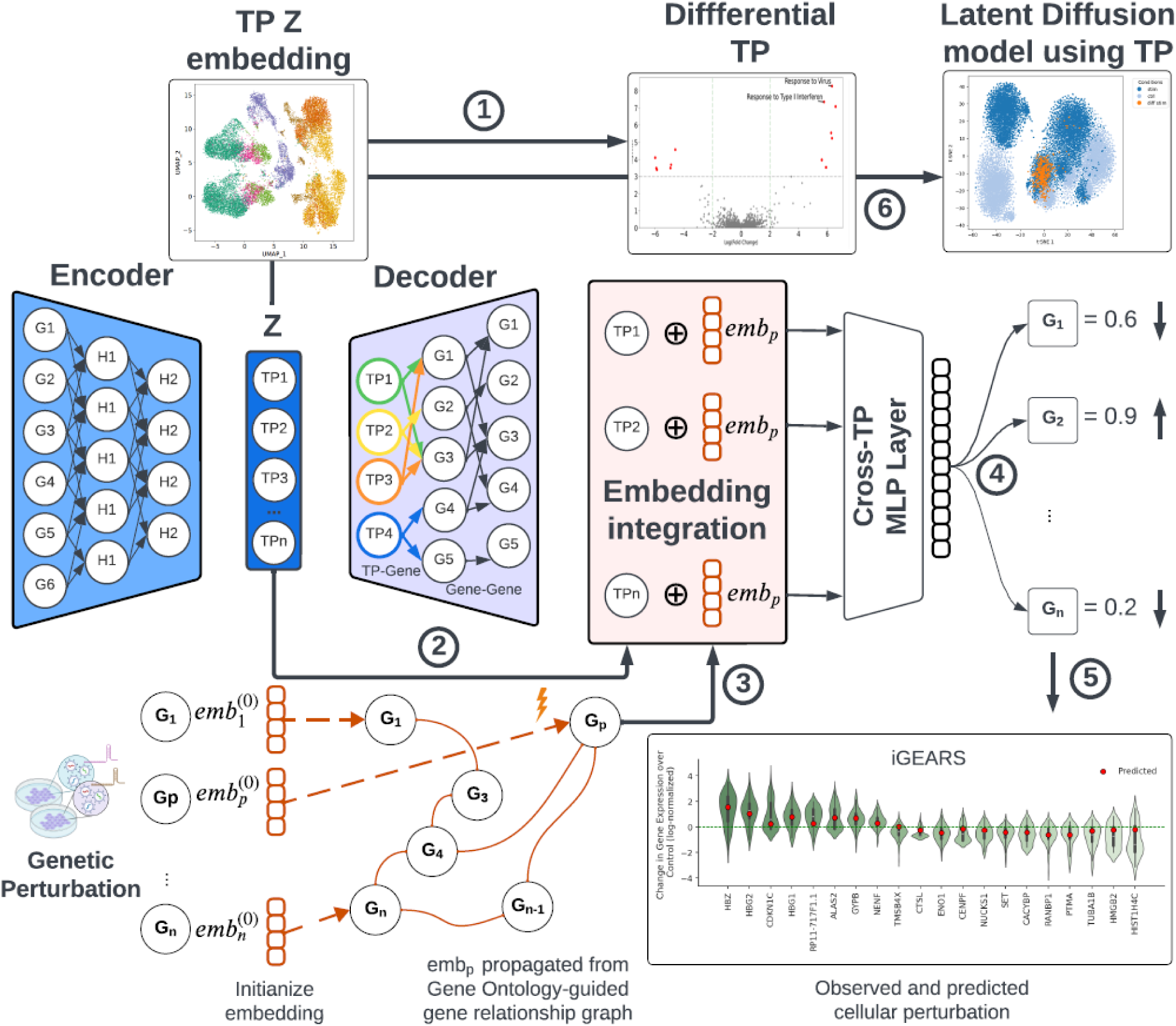
Overview of iGTP framework: The single-cell data is processed by the iGTP non-linear Encoder, generating the transcriptional program (TP) embedding (z) using reparameterization for input TPs. The Decoder consists of two layers: 1) The TP-Gene layer, 2) The Gene-Gene layer. Both layers apply a sparse matrix to the gradient to eliminate non-existent connections, leaving only the true connections. For instance, in the TP-Gene layer, the value is one if a gene is present in a given TP; otherwise, it’s zero. During backpropagation, the weight of the true connection is retained, while all others are forced to zero. The two layers are used to reconstruct the gene expression within each cell, allowing each TP to be learned by TP-related genes and the gene-gene interaction within those genes. The TP embedding ***Z*** and gene layer is then extracted for visualization and differential TP analysis (step 1). Additionally, TP embedding ***Z*** (step 2) can be concatenated with gene perturbation embeddings (emb_p_) derived from one PPI-guided gene relationship graph (step 3). This graph is generated using genetic perturbation data and graph propagation of the genetic perturbation impact *(P* on gene *G_p_*), constructed from gene-gene relationship derived from gene ontology (GO). The concatenated embeddings are then processed through a two-layer Multilayer Perceptron (MLP), designed to associate TPs with genes. The results in the model outputting altered gene expressions (step 4) represented the predicted overall impact of perturbations on other genes. The top altered gene expressions were ordered and highlighted using a boxplot (step 5). Finally, the TP embedding ***Z*** could be used for latent diffusion model to generate cells for given cell types and condition (step 6).

### Distilling biological insights with interpretable latent space framework

We first evaluate the capacity of the iGTP model to unveil the ground-truth biological processes in interferon-β-treated Peripheral Blood Mononuclear Cells (PBMC) dataset^17^. We input the single-cell dataset and enforce iGTP to learn the gene ontology (GO) as the TP and PC commons as the PPIs. After training, we reparametrized the latent space ***Z*** generated from iGTP on 2D t-distributed stochastic neighbor embedding (TSNE). As shown in Fig. 2a & 2b, iGTP differentiated both cell types and stimulation states. In Fig. 2c, we found that the TP “GO:0035456: response to interferon-β” simulation had overall higher activities in stimulated cells, confirming the ability of iGTP to capture pathway activity in its latent space. Comparable cellular “GO:0035455: response to interferon-α” and “GO:0035456: response to interferon-β” were observed in immune cells^18^. The iGTP framework captured the higher PPI activity between ISH15 and IFNB1 (Interferon Beta 1) within the BP TP of “GO:0035456: response to interferon-β” in the stimulated cells (Fig. 2d). We also benchmarked against the conventional single sample Gene Set Enrichment Analysis (ssGSEA) method^19,20^ to calculate the activity of the same TPs in each cell. As shown in Fig. 2e, the TP activity of response to interferon-β is much lower (or close to resting status) in stimulated adaptive immune cells (CD8+ T cells, CD4+ T cells, and B cells) than in other stimulated innate immune cells (NK cells, Dendritic cells, FCGR3A+ monocytes, and CD14+ monocytes). Compared to Fig. 2c & 2e, both algorithms exhibited similar patterns, but iGTP demonstrated a stronger change in biological activity. The phenomenon was further validated by a recently published paper^18^, suggesting ssGSEA and iGTP could capture the response to interferon-β activity. As shown in Fig. 2f, the volcano plot for differential TPs across all cell types between stimulated vs control with plaid-based Bayers factor as the fold changes magnitude (Table S1). The top stimulated TPs were BP of “GO:0009615: response to virus”, BP of “GO:0034340: response to type I interferon”, and other interferon-related TPs. We also benchmarked the conventional gene-set enrichment analysis (GSEA) method to measure the activity of the same TPs in each cell type with iGTP results. In Fig. 2g, we showed the TPs in both Bayers factor from iGTP and normalized enrichment score from GSEA. We pinpointed that BP of “GO:0009615: response to virus” and other interferon-related TPs were in the top-right corners of CD8+ T cells (Table S2) and CD14+ monocytes (Table S3). Interestingly, the BP TP of “GO:0045088: regulation of innate immune response” was predicted to be much lower in CD8+ T cells than CD14+ monocytes by iGTP. On the other hand, GSEA predicted it among the top TPs in both cell types, which is contradicted by the fact that TP “regulation of innate immune response” is mainly regulated in innate immune cells instead of CD8+ T cells^21^. Collectively, these findings imply that iGTP TP accurately represents the anticipated primary biological pathways in PBMC, thereby potentially serving as a valuable tool for projecting cells from other datasets into an interpretable framework. This approach enables the detailed exploration of cell-type-specific patterns at the level of cellular processes. Lastly, we leveraged this PBMC dataset to comprehensively compare our iGTP model and another deep learning model VEGA, with different knowledge of PPI and TP databases. Then, we evaluated the model performance using NMI, ARI, silhouette metrics from scvi-tools^22^, and graph connectivity for evaluation. Overall, iGTP (GO TP+PPI compiled) outperforms ssGSEA and VEGA, as evidenced by the average of all evaluation metrics (Fig. 2h).

**Fig. 2.**
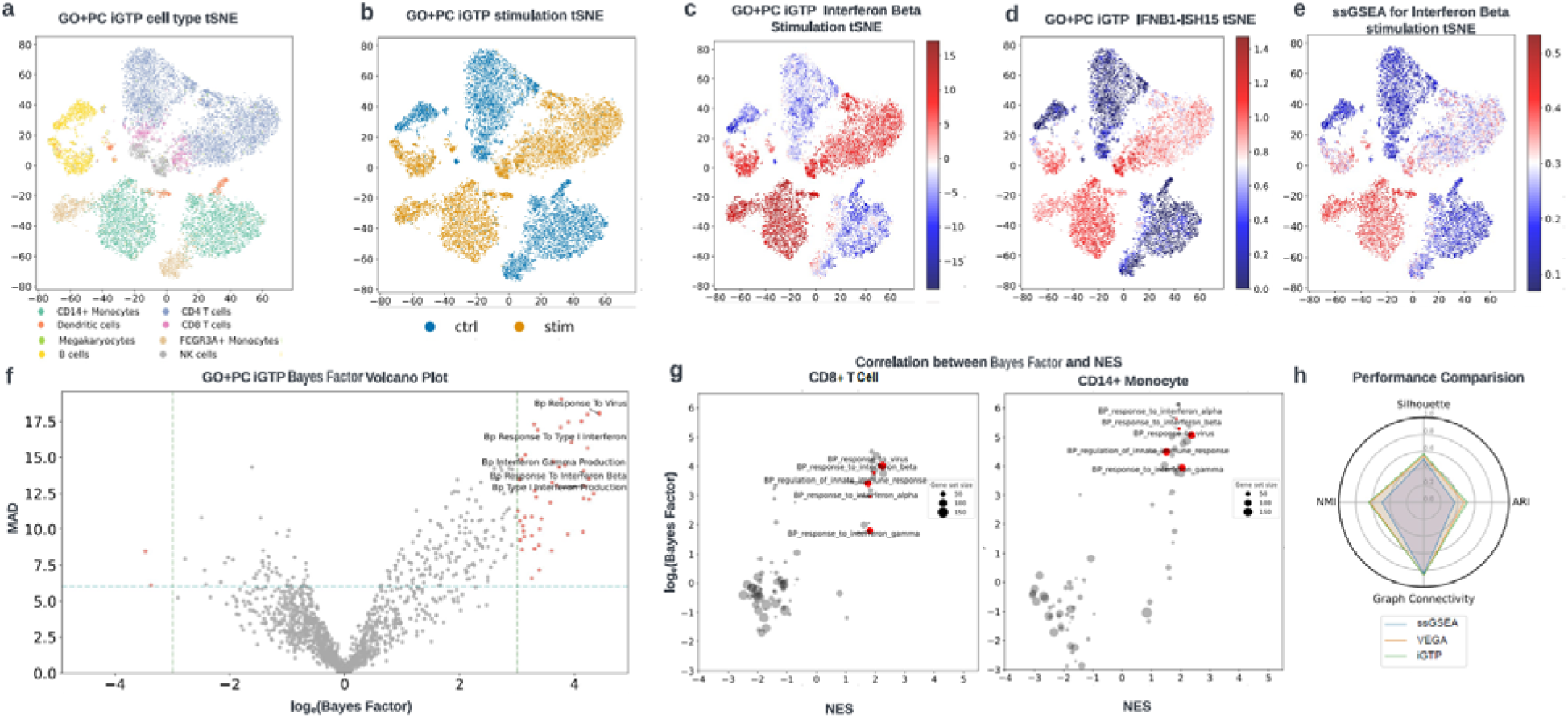
Benchmarking the model performance of iGTP on stimulated and resting PBMC dataset. First, we evaluate the ability of iGTP to distinguish between different cell types and stimulation states. Panel a. labeled the cells on the t-SNE plot based on their cell types, while panel b. labeled them based on the stimulation condition. c. The score indicated the activity of interferon beta in each cell. Higher activity (red) was observed in interferon beta-stimulated cells compared to the unstimulated cells (blue). NK cells, monocytes, and dendritic cells with higher biological activity than other cells. d. We labeled the PPI score (ISH15 and IFNAR1) on the same map and find that stimulated T cells (CD4+ and CD8+) have lower activity than stimulated monocytes. Panel e indicates that the conventional ssGESA results are similar to the iGTP results for the TP (panel c). f, The featured biological pathways (|log_e_(Bayes factor) > 3 & mean absolute difference (MAD) > 6) were highlighted in the volcano plot. Additionally, a positive ln (Bayes factor) indicates a Transcriptional program (TP) with higher activity in stimulated cells than unstimulated cells. A higher MAD indicates a more significant difference within the latent space between stimulated and unstimulated cells. g. The correlation between the Bayes factor and NES (GSEA) was compared in two different cell types, CD8+ T cells and CD14+ monocytes, focusing on highly differentiated TPs based on the Bayes factor results. h. Several state-of-the-art methods, including ssGESA, VEGA, and iGTP, were benchmarked using performance metrics from scvi-tools.

### iGTP model captures both known and unknown TP activities related to brain cellular function and Alzheimer’s disease

To explore the generalizability of our iGTP model, we analyzed brain cells from Alzheimer’s disease (AD) and cognitively normal (CN) individuals who share similar AD risk backgrounds. We adapted AD and brain function-related TP curation from our previous work^23^. The normalized mean TP activities by each cell type and disease condition were visualized in Fig. 3a.

**Fig. 3.**
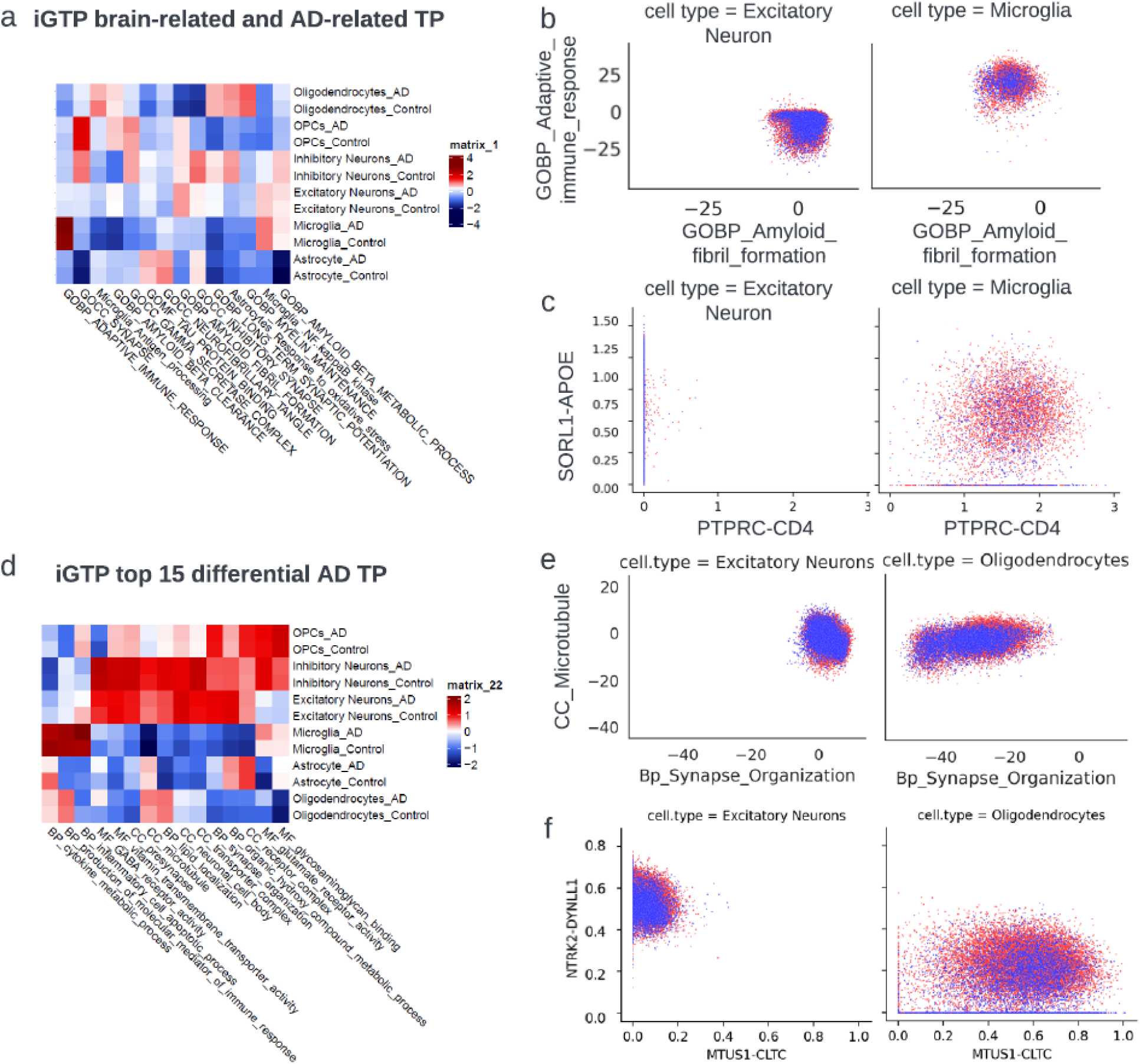
TP embeddings characterize biological activities of brain-related and AD-related functions. a. Heatmap for visualizing the mean TP embedding values in each cell type and condition for 14 representative TPs for brain-related and AD-related functions. The mean TP embedding values were conducted z-score transformation within each TP. b. Dot plot visualizing the cellular relationship in TP embeddings between two TPs across two cell types, stratified by AD in red and CN groups in blue. c. Dot plot visualizing the cellular relationship in TP embeddings between two PPI pairs across two cell types, stratified by AD (red) and CN (blue) groups. d. Heatmap for visualizing the mean TP embedding values in each cell type and condition for the top 15 most differential TPs in AD and CN from GO TP sets. The mean TP embedding values underwent a z-score transformation within each TP. e. Dot plot visualizing the cellular relationship in TP embedding values between two top TPs across two associated cell types, stratified by AD (red) and CN (blue) groups. f. Dot plot visualizing the cellular relationship in TP embeddings between two TP PPI pairs across two associated cell types, stratified by AD (red) and CN (blue) groups.

We identified the TP activities that were enriched with corresponding cell types, such as BP of “adaptive immune response in Microglia”, cellular component (CC) of Synapse in both inhibitory neurons and excitatory neurons, suggesting TP embedding from our iGTP model could capture meaningful biological activities. Moreover, we also identified a few well-known AD-related TP with more activated states, such as molecular function (MF) of “GO:0048156: Tau protein binding” in AD excitatory neurons^24,25^ and BP of “GO:1990000: Amyloid fibril formation” in AD excitatory and inhibitory neurons^26^. The embeddings of these TPs were further visualized in 2D (Fig. 3b) for astrocyte and microglia. The dot shifts between AD and CN indicate our TP embeddings indicate biological differences in AD and CN. More interestingly, we delineated PPI connections, PTPRC-CD4^27^ and SORL1-APOE, linked to BP “GO:0002250: Adaptive immune response” and BP of “GO:1990000: Amyloid fibril formation” respectively, across cell types. Fig. 3c illustrates the cell-type-specific PPI activities in excitatory neurons and microglia between AD and CN cells. To identify potential unknown GO TPs that predominantly distinguish AD from CN by iGTP, we highlighted the 15 most variable TPs (Fig. 3d), including a few neurotransmitter receptors (such as MF of “GO:0008066: glutamate receptor activity”, MF of “GO:0016917: GABA receptor activity”, and MF of “GO:0005539: glycosaminoglycan binding”) have higher activities in excitatory neurons, inhibitory neurons, and OPCs. MHC protein complex was mainly expressed in Microglia, OPCs, and Oligodendrocytes. The corresponding TP activities from ssGSEA (Fig. S1) exhibit a median Spearman correlation coefficient *ρ* of 0.74 across cell types for these TPs. As shown in Fig. 3e, the difference in cellular embedding distribution between AD and CN indicates our TP embeddings in excitatory neurons and microglia in synapse-related functions (BP of “GO:0050808: synapse organization” and CC of “GO:0005874: microtubule”).

In our analysis of these TPs, we specifically identified PPIs with significant differences between AD and CN cells. For example, the NTRK2-DYNLL1 link in the BP of synapse organization showed a notable disparity, where a variant in NTRK2 has been previously associated with AD in earlier research^28^. As Fig. 3f illustrates, the PPI between MTUS1-CLTC in the BP of microtubule formation exhibits distinct cellular activities in AD compared to CN individuals. This observation is consistent with previous studies showing elevated MTUS1 expression in specific brain regions of AD patients relative to CN controls^29^.

### Generalizability to predict out-of-sample single-cell PBMC data

Next, we examined if iGPT could generalize to accurately infer an interpretable latent representation of data not seen during training (out-of-sample data). We evaluated iGTP in two settings for this purpose. In the first case, we assessed the biological generalization of iGTP’s inference by excluding some pairs from the same dataset during training. Specifically, we explored whether the inferred iGTP activities for the omitted cells conveyed meaningful biological information while not in training. For this, we excluded cells infected by influenza from the COVID-19 dataset during training. Then, we inferred the PBMC “GO:0009615: response to virus”-related TP activities on the removed samples (influenza) using the model trained by the remaining cells. As shown in Fig. 4a, CD14+ monocytes and dendritic cells from severe influenza patients tend to have higher activities than cells from COVID-19 Low-Dose Naltrexone (LDN) treatment or healthy controls in BP TP of “GO:0034340: response to type 1 interferon” and TP “GO:0009615: response to virus”. We selected the 6 interferon-related TPs (Table S4) and 4806 related protein-protein interactions (PPIs) which represent the biological activities of virus invasion across various cell types. We tested the Pearson correlation coefficient *r* of the Bayes factor in pretrained samples and predicted samples. All cell types, except for B cells, have an intra-sample correlation of *r* > 0.4 (Fig. 4b & 4c). Next, we explored whether the iGTP model could predict this unseen similar cellular condition population from a new batch study. Here, we trained our model using the COVID-19 dataset and predicted the TP activities in the Kang et al. dataset. As shown in Fig. 4d, CD14+ monocytes and dendritic cells from the Kang et al. dataset tend to have higher activities than in interferon-β stimulated cells and lower activities in resting cells from COVID-19 dataset in BP TP of “GO:0034340: response to type 1 interferon” and TP “GO:0009615: response to virus”. We further tested the coefficient r of the Bayes factor between the COVID-19 dataset and predicted the Kang et al. dataset by each cell type. The overall prediction performance in Fig. 4e & 4f failed to reach the performance in Fig. 4b & 4c, suggesting the virus infection conditions on heterogeneous individuals have a more complex impact than the cellular response to stimulation of interferon-β only^30,31^.

**Fig 4.**
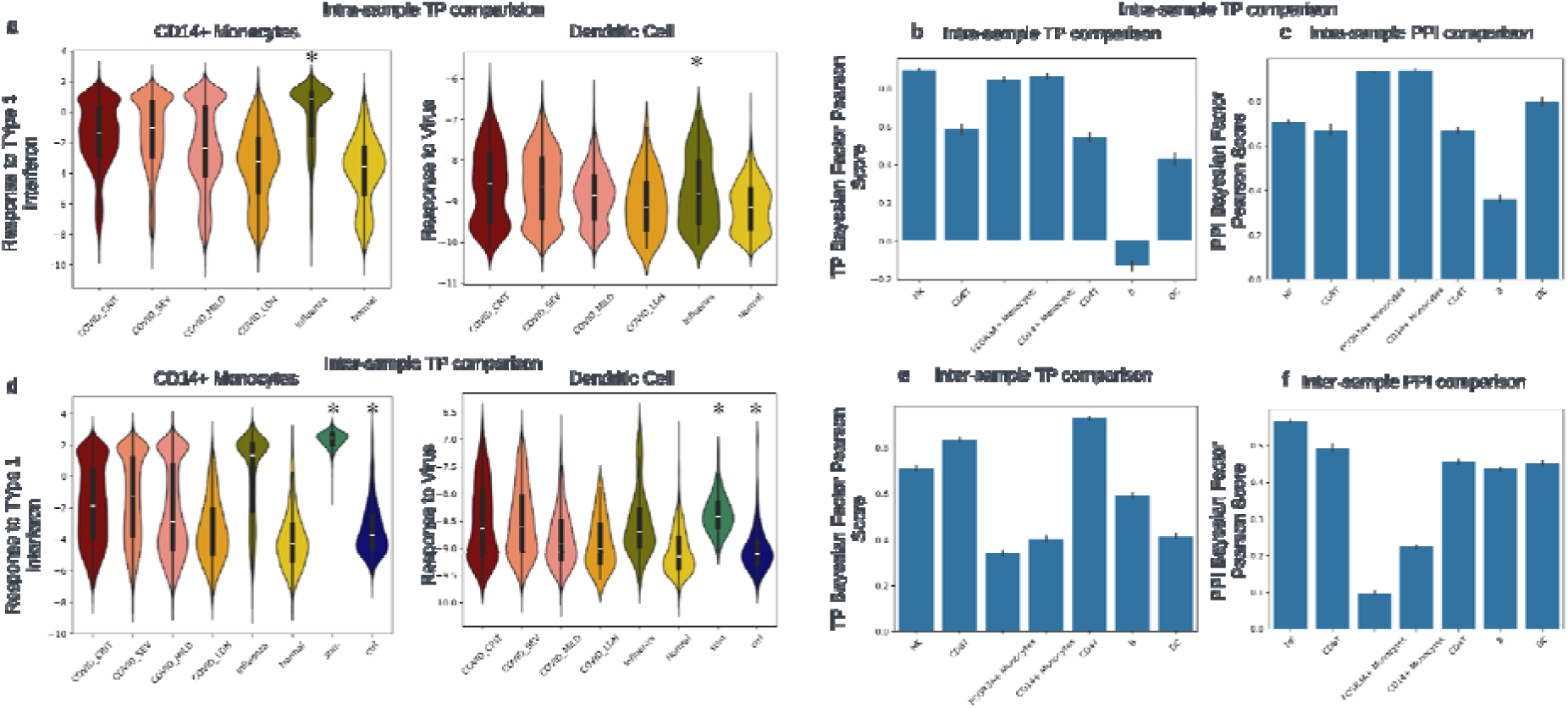
Prediction performance of intra- and inter-sample for representative TPs and PPIs. a. Distribution of TP activities in COVID-19 PBMC training samples, alongside predictions of intra-sample activities in the testing dataset. Specifically, the sample source labeled with an asterisk (*) represents the unseen intra-sample (testing sample) from the same COVID-19 PBMC dataset. For this testing sample, TP activity predictions were made using a pretrained model, which was trained on all other cell types. Two TPs were used to demonstrate the related biological mechanism in CD14+ monocytes and dendritic cells, respectively. b. Bar plot for each cell type shows the distribution of Pearson correlation coefficient *r* between the Bayes factor (BF) of selected TPs from GO in pretrained samples and the corresponding predicted TP BF values for the unseen intra-samples (testing sample) on the same COVID-19 PBMC dataset. c. Bar plot for each cell type shows the distribution of Pearson correlation coefficient *r* between the PPIs of the selected 6 interferon-related TPs from GO in pretrained samples and the corresponding predicted PPI BF values for the unseen intra-sample (testing sample) on the same COVID-19 PBMC dataset. d. Distribution of TP activities in COVID-19 PBMC training samples, alongside predictions of inter-sample activities in the Kang et al. PBMC dataset. Specifically, the sample source labeled with an asterisk (*) represents the unseen intra-sample (testing sample) from Kang et al. PBMC dataset. For this testing sample, TP activity predictions were made using a pretrained model, which was trained on all cell types from COVID-19 PBMC dataset. Two TPs were used to demonstrate the related biological mechanism in CD14+ monocytes and dendritic cells, respectively. e. Bar plot for each cell type shows the distribution of Pearson correlation coefficient *r* between the BF of selected TPs from GO in pretrained samples and the corresponding predicted TP BF values for the unseen inter-sample (testing sample) of the Kang et al. PBMC dataset. f. Bar plot for each cell type shows the distribution of Pearson correlation coefficient *r* between the PPIs of the selected 6 interferon-related TPs from GO in pretrained sample and the corresponding predicted PPI BF values for the unseen intra-sample (testing sample) of the Kang et al. PBMC dataset.

### Inference unseen perturbation on genes using pretrained TP embeddings

Next, we investigated if our distinct TP embedding and Decoder architecture could effectively capture the impact of perturbations on gene expression levels, thereby offering *in silico* assessment of the perturbation response. Consequently, we have integrated our TP embedding with a perturbation relationship graph, which is an adaptation from GEARS, a deep learning framework based on Graph Neural Network (GNN) architecture^16^. The Norman et al. dataset^32^ contained 105 single CRISPRa perturbations on K562 chronic myelogenous leukemia cell line. As shown in Fig. 5a, we input the gene matrix of unperturbed K562 cells in the same study to obtain the unperturbed TP embeddings. We combined TP embedding with knockdown gene embedding generated from the perturbation graph, where edges connecting genes are extracted from GO (Fig. 1). After two layers of GNN training, gene embedding representing its spatial pattern inherited in the perturbation graph further concatenates TP embedding as the input of perturbation prediction layers. As shown in Fig. 5a, we predicted the mean difference after perturbation of MAPK1 and compared the top 20 most differentially expressed genes with the real perturbation distribution. We benchmarked our iGTP framework with GEARS and identified a similar overlap (10 out of 20 genes) and trend of the top 20 most differentially expressed genes (Fig. 5b). Lastly, we observed a much larger perturbation in genes within perturbed TP than in genes out of perturbed TP with t-test p-value 1.97^-72^ and Mann-Whitney U test p-value 1.24^-76^ (Fig. 5c).

**Fig 5.**
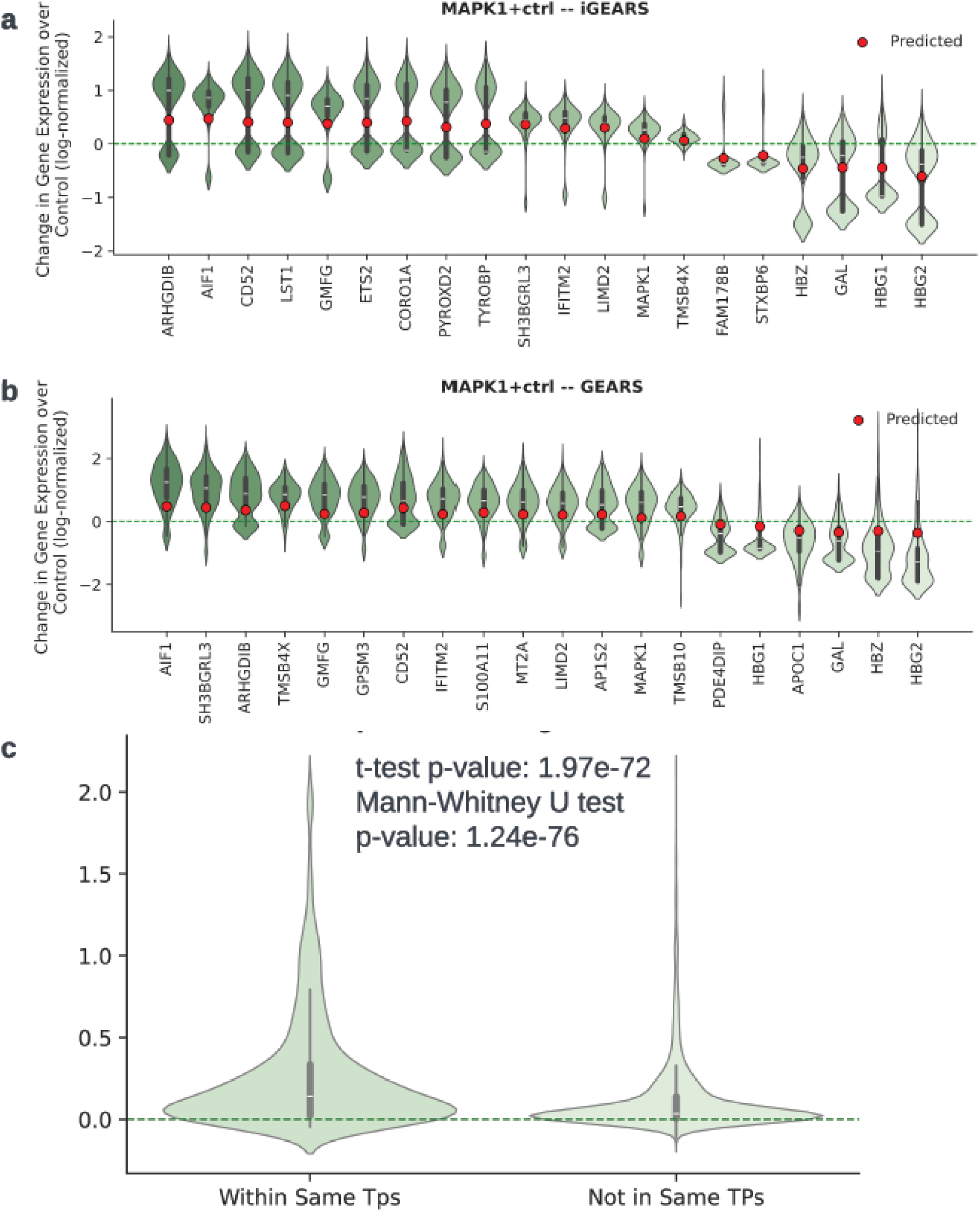
**The Comparison between GEARS and iGEARS on the perturbation prediction** iGTP perturbation (iGEARs) assesses the change in gene expression and benchmark with GEARs. a&b, Changes in gene expression in iGEARs and GEARs. Violin plot and Boxplot showed experimentally measured gene expression difference after perturbing the corresponding gene, where top 20 post-perturbation genes in response to perturbation. Genes were ordered by change magnitude. The red dots show the mean change in gene expression predicted by our model. The green dotted line represents unperturbed control gene expression. a&b showed top 20 variated genes after perturbation on gene MAPK1 in iGTP. c. We investigated the change in gene expression over MAPK1 perturbation. We tested whether genes in the TP group containing MAPK1 or genes in the TP group excluding MAPK1 differed significantly, using t-tests and Mann-Whitney U-tests to compare their mean differences.

### SHAP importance of TPs in predicting PBMC states and cell-type-specific PPIs

We examined the performance of iGTP and the relationship between the layers of the Decoder. Initially, we evaluated if the iGTP embedding can differentiate the stimulation state in the PBMC dataset from Kang et al (Fig. 6a). We used the embeddings of iGTP TP, iGTP PPI, and VEGA, as well as conventional ssGSEA and raw gene expression features to measure the AUC for PBMC labels. iGTP TP embedding had the highest AUC (0.9904) in our comparison. We then utilized SHapley Additive exPlanations (SHAP) values to validate the contributions of TP embeddings in differentiating stimulation state. Fig. 6b presents the SHAP values for the top 10 TPs and the aggregate value for the remaining TPs. Consistent with expectations, many of the highest-ranking TPs closely corresponded with those exhibiting the highest Bayes factors (Fig. 2f). Furthermore, TPs associated with shared functions, such as BP “GO:0009615: response to virus” and BP “GO:0043331: response to dsRNA”, exhibited similar SHAP value distributions. In Fig. 6c & 6d, we further dissected the PPI embeddings in these two TPs among different cell types. Specifically, we selected the top 10 PPIs with the largest MAD differences between stimulation and control statuses from those shared by CD8+ T cells and CD14+ monocytes. Interestingly, we found that the BST2-TRIM25 PPI within the “GO:0035455: response to interferon-α” TP had the highest difference. The interferon-inducible protein, BST-2 (or, tetherin), plays an important role in adaptive immune response^33^ and TRIM25, also known as tripartite motif-containing 25, can regulate the immune response of viral infection^34^. Absolute MAD has a higher value of this PPI in CD8+ T cells than CD14+ monocytes. On the other hand, the RSAD2-APOC1 PPI within the “response to type one interferon” TP was the second most differentiable between CD8+ T cells and CD14 + monocytes (Fig. 6d). Recent research indicates that RSAD2 is up-regulated when exposed to type one interferon at the monocyte’s single-cell level^18^. These findings suggest that our iGTP model could capture the top TPs and fine-grained cell-type-specific responses PPIs response to interferon-β stimulation. To delineate the relationship between layers of the iGTP Decoder, Sankey Diagram (Fig. 6e) shows the relative weight flow, in between TP “GO:0035455: response to interferon-α” to IFITM3 and GATA3 genes within the TP. Intuitively, the thickness of the link between TP and PPIs reflected the normalized importance of PPIs to the TP. The PPI IFITM3-STAT3 was found to have the largest contribution to this TP, with the IFITM3/STAT3 axis playing a key role in regulating immune responses through interferon signaling. IFITM3 modulates the phosphorylation and activation of STAT3, thereby influencing gene expression related to inflammation and immunity^35^. Similarly, the thickness between the PPIs and genes suggested the relative importance of gene to this PPI. The final layer indicated the relative importance of genes within this TP, with gene IFITM3 making the largest contribution.

**Fig 6.**
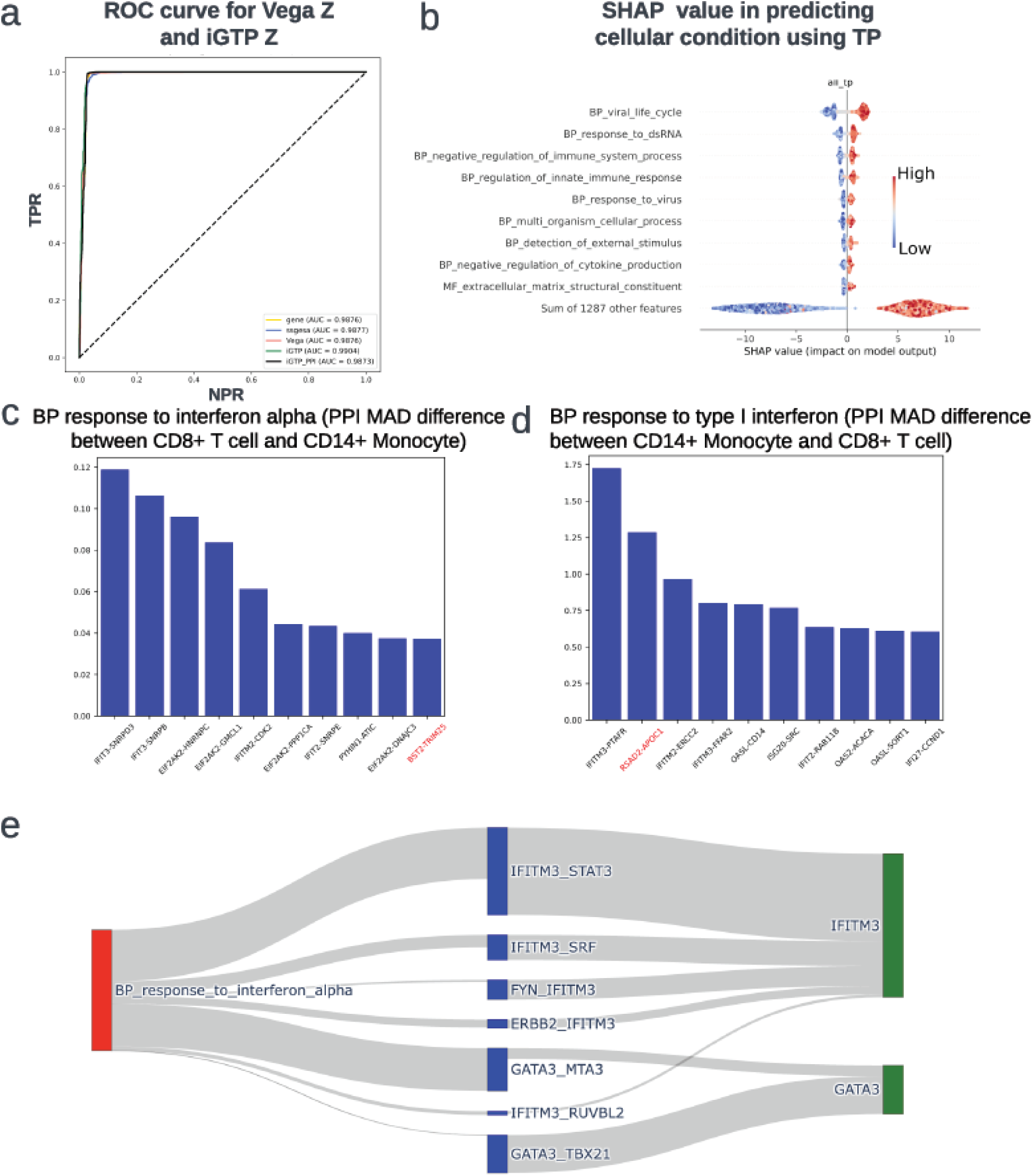
Model performance and interpretability for TP, PPI, and gene on Kang et al. PBMC dataset. a. Benchmark prediction as stimulated cell label on Kang et al. PBMC dataset. b. SHAP value derived from XGBoost, utilizing all TP embedding z values as the features (X) and the cellular state (either ’stimulated’ or ’control’) as the label (Y). Beeswarm plot for the top 10 TPs with the biggest absolute SHAP values and the sum of all other features. c & d, Barplot for top 10 MAD difference of PPIs derived from stim vs ctrl states in CD14+ monocyte and CD8+ T cell for “GO:0035455: response to interferon-a” and “GO:0034340: response to type I interferon”, respectively. e. Sankey diagram illustrated relative weight flow in Decoder of iGTP framework for TP “GO:0035455: response to interferon-a” and their related genes within TP, showing how TP embeddings towards reconstruction of gene expression levels through PPIs and relative gene importance from certain TP.

### Conditional generation of new cells using diffusion model and TP embeddings

The denoising diffusion probabilistic models (DDPM)^36^, widely used in image generation, starts with random noise and iteratively refines it into a coherent image by reversing a diffusion process through a series of conditionally learned denoising steps. To further explore the generative application of our iGTP model, we revised and integrate one recent single-cell latent diffusion model scDiffusion^37^ into our iGTP framework and use iGTP TP embedding as the latent input Xo. To develop a robust pretrained model, we utilized the previous Ahern et al.^38^ COVID-19 PBMC dataset, which contains over 600,000 cells, to pretrain the iGTP model with non-redundant GO TPs. Then, we input Kang et al.^17^ dataset through this pretrained encoder to obtain the TP embeddings (X_0_) of ∼13,000 PBMC cells with interferon-beta stimulation and control. As shown in Fig. 7a&7b, most diffusion-generated control CD4+ T cells were located in the overlapping region of control state cells and CD4+ T cells, suggesting the potential of conditionally generating data on specific cell types and states. We designed a classifier to distinguish control CD4+ T cells (including both diffusion-generated and original) from other cell types, using TP embeddings as features. This approach achieved an accuracy of 0.98 on the test set (Fig. 7c) and an area under the precision-recall curve (AUPRC) of 0.98 ± 0.00 in five­fold cross-validation (Fig. 7d).

**Fig 7.**
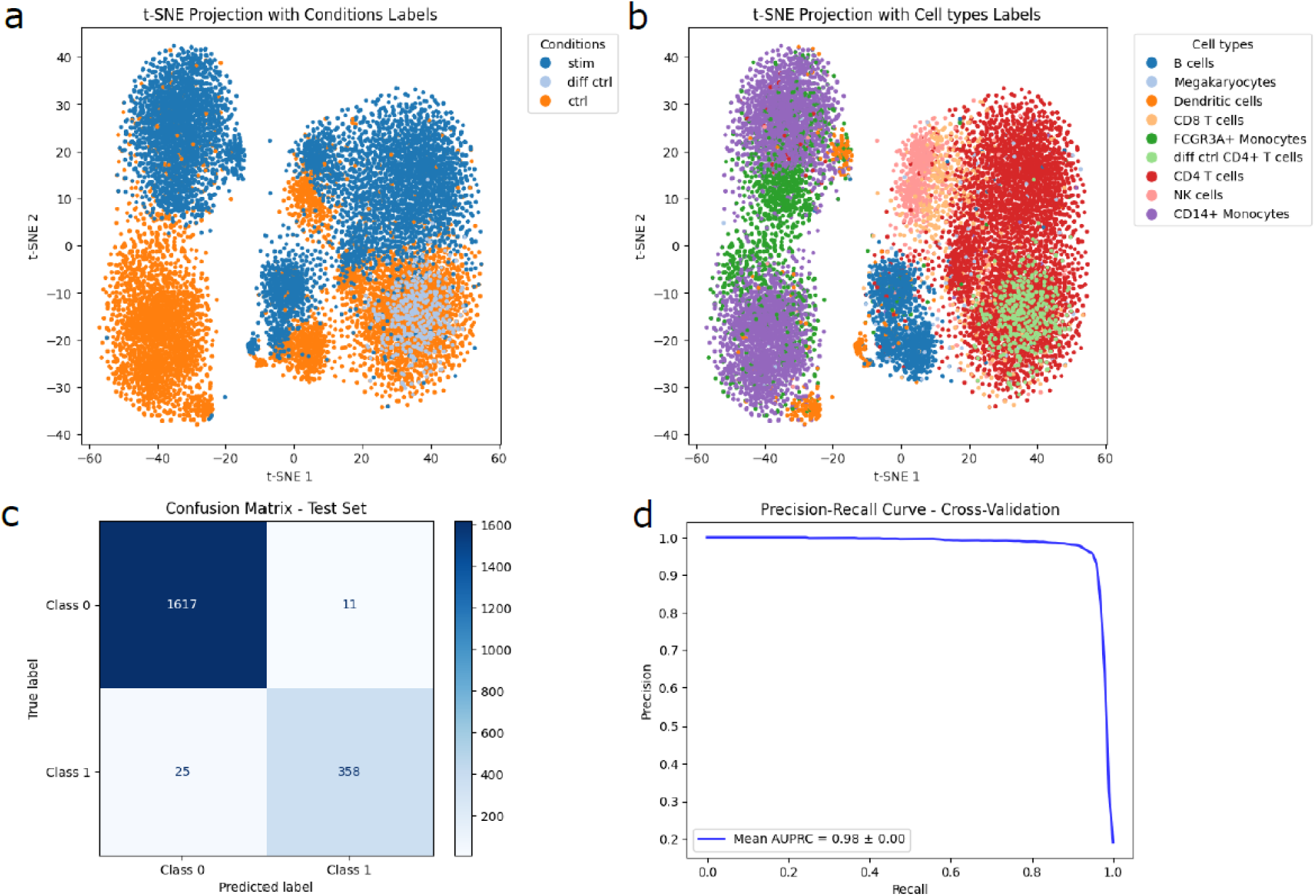
Latent Diffusion Model to generate given cell type and condition using TP embeddings on Kang et al. PBMC dataset. a. t-SNE embedding for PMBC datasets and diffusion generated control CD4+ T cells by conditions. b. t-SNE embedding for PMBC datasets and diffusion generated control CD4+ T cells by cell types. c. Confusion matrix illustrating the prediction performance on the test set of PBMC datasets. In this analysis, diffusion-generated control CD4+ T cells and actual control CD4+ T cells were labeled as 1, while resting cells were labeled as 0. d. The AUPRC plot evaluating the performance of predicting control CD4+ T cells using 5-fold cross-validation.

## Discussion

The deep-learning framework VAE has gained more attention for the scalability and flexibility to handle the growing single-cell RNA-seq data. In this work, we developed an interpretable generative transcriptional program (iGTP) VAE-based framework that quantifies the biological networks composed of TPs and PPIs in driving the cellular state alteration. We used examples in the PMBC dataset with interferon-β treatment dataset, COVID-19 PMBC dataset, and AD dataset, suggesting our iGTP model could accurately capture the known biological process and discover new TP that was not well-characterized before. Moreover, we tested our model robustness within intra-sample and inter-sample tasks, showing the VAE has the potential transferability. Lastly, we integrated one GNN-based module to provide *in silico* perturbation estimation for the known perturbation effect. The integration of this feature substantially augments our model’s proficiency in accurately interpolating responses to perturbations at each TP. This advancement demonstrates substantial potential in facilitating extensive *in silico* predictions, which could be pivotal for strategizing and optimizing real-world experimental approaches. Finally, we demonstrated an application of our iGTP TP embeddings by integrating iGTP with a latent diffusion model, successfully generating accurate new cell embeddings for specified cell types and states.

In this study, we used gene expression to approximate protein, PPI, and TP level activities. We innovatively designed a biologically meaningful Decoder that integrates prior knowledge, enabling the VAE model to capture the hierarchical structures of biological processes among TPs, PPIs, and genes. In the Kang et al PBMC dataset, we successfully replicated the BP of “GO:0009615: response to virus”, BP of “GO:0034340: response to type I interferon” as the biological processes corresponding to interferon-β treatment (Fig. 2f & 6b). In addition, the model not only differentiates the cell state (w/wo interferon-β stimulation) but also captures the activity difference between monocytes and T cells (Fig. 2c). Moreover, we identified several key PPIs with cell-type-specific importance, such as BST2-TRIM25 and RSAD2-APOC1, which exhibited the highest MAD differences between stimulated and control conditions for CD4+ T cells and CD14+ monocytes (Fig. 6c & 6d). This indicates that these PPIs within the TPs contribute significantly to the alteration of cellular states. In the AD dataset, we confirmed iGTP could capture the cell-type-specific TP activities and pinpointed a few novel TPs might be responsible for the molecular mechanism for AD pathogenesis. Even though we obtained a Spearman correlation coefficient *ρ* of 0.74, some top differential TP was predicted to flip in cells from AD and CN individuals between the results of iGTP and ssGSEA. For example, iGTP identified the higher activity of CC “GO:0005874: microtubule” in synapse, oligodendrocytes, and astrocytes for cells from AD individuals, while the activity signals were predominant in synapse only in the result of ssGSEA (Fig. S1). In oligodendrocytes, microtubules play a crucial role in extending processes to contact axons and elongating the myelin sheath. In astrocytes, neighboring cells and the extracellular environment can influence changes in cell shape and branching^39^. The BP “GO:0050808: synapse organization” is essential for neuron communication. In Alzheimer’s, amyloid plaques and tau tangles disrupt this, leading to cognitive decline. OPCs play a role in signal transmission by interacting with neurons and forming neuronal-OPC synapses, which exhibit synaptic plasticity like that observed in neuron-neuron synapses^40^. However, while iGTP identifies signals in OPCs, ssGSEA fails to detect much activity (see Fig. S1). This discrepancy might be raised by the methodology difference. As ssGSEA^19,20^ calculates each cellular TP score by comparing the distribution of gene expression ranks inside and outside the TP, it may fail to capture the complex, non-linear relationships that iGTP can. Therefore, further analysis incorporating ground truth knowledge about TP activities is warranted.

We evaluated the transferability of iGTP on PBMC datasets subjected to different immune stimulations: one from a virus (COVID-19 and influenza) and the other from interferon-β. Viral invasion can stimulate interferon-β production, boosting the immune response in a similar way to interferon-β stimulation. The model was trained on COVID-19 patients in various conditions, including critical, severe, mild, receiving COVID-19 treatment, and normal. It can predict the biological activity in two TPs: the BP of “GO:0009615: response to virus” and the BP of “GO:0034340: response to type I interferon” in previously unseen patients with severe influenza infection. The model also shows the difference in biological activity linked to the conditions (Fig. 4a). When the entire COVID-19 dataset was used to train iGTP and predict the Kang et al. PBMC dataset, the model could differentiate the cell state with and without interferon-β stimulation (Fig. 4d). Both results suggest that iGTP can transfer knowledge from one dataset to another while maintaining some common biological rationale. However, overall prediction performance in the same dataset is better than in different datasets (Fig. 4b, 4c, 4e & 4f). This suggests that virus infection conditions in diverse individuals have a more complex impact than the cellular response to stimulation of interferon-β alone.

As shown in Fig.6a, our iGTP model obtained a very close AUROC among conventional bioinformatics approaches and other deep-learning-based methods in predicting PBMC data with stimulation or not. Compared to traditional bioinformatics tools, deep learning-based methods demonstrate significant advantages in learning feature representation within high­dimensional spaces. These vectorized spaces could be further integrated with other frameworks (such as GEARS) to handle a more complex task, such as response to perturbation prediction. Although the reconstruction rate of iGTP is lower compared to the VEGA model, particularly against designs using fully connected layers or a single sparse masker layer (as employed by VEGA), this trade-off is intentional to enhance the interpretability of biological mechanisms in our approach. We envision GEAR in iGTP, which could also be useful to prioritize drugs based on pathway expression in cancer, as studying the response of specific cell populations may inform drug sensitivity and resistance. Integrating drug response prediction models with such explanatory models could benefit designing novel therapeutic strategies.

## Methods and Materials

### iGTP framework

iGTP is a deep learning framework based on the Variational AutoEncoder (VAE) structure and utilizes an Encoder with fully connected layers to process input gene expression data (***X***). We transformed gene expression data into a biologically interpretable latent space to approximate the activities of protein-protein interaction (PPI), and biological pathway or transcriptional program (TP). From the TP space, a vector ***Z*** undergoes decoding through a specifically designed network to reconstruct the gene features ***X***.

iGTP differs from the traditional VAE model in that the Decoder network is designed to incorporate prior knowledge of the relationship between TP and genes. Specifically, we employ a sequence of masked fully connected layers as the Decoder network to reconstruct the gene features ***X***.

The vector of gene features ***G*** *= {g_1_,g_2_,,…,g_NGene_*} were encoded to derive latent variables ***Z*** = {z_1,_…,z*_NTP_*}. Each latent variable of ***Z*** corresponds to a single transcriptome program. To ensure that each dimension of ***Z*** is a TP (scalar), we enforce the Decoder network to embed only (masked) linear operations, i.e. masked fully connected layers.

To reconstruct the gene features, ***Z*** is transformed by a Decoder network with two masked fully connected layers. First, ***Z*** is transferred into gene by going through the first masked fully connected layer, whose weights are a binary matrix *W^TP-Gene^.* The weights of *W^TP-Gene^* are predefined using prior knowledge of TP-gene relations: for each entry *W^TP-Gene^(i,j),* its value is one if the í-th gene is a member of j-th TP, and zero otherwise. Second, the output matrix, denoted as ***O***, will go through another masked fully connected layer, *W^TP-Gene^*, to reconstruct the gene features *G.* Each entry of *W^Gene-Gene^* (*i,j*) denotes whether the i-th gene and *j-th* gene has PPI relations: one if the relation exist and zero otherwise. The weight matrix *W^Gene-Gene^* is, therefore, a symmetric matrix. The Decoder network reconstructs gene feature vector ***G*** = {*g_1_ g_2_*, …., *g_NGene_*} as following:

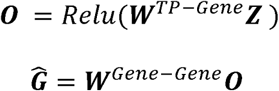

Like the traditional VAE, the objective of training iGTP is to reconstruct gene expression ***X*** from the TP space ***Z*** by maximizing the reconstruction likelihood,

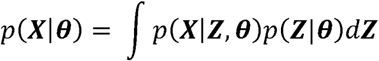

where *θ* is the parameters of iGTP, ***X*** is the input gene expression data, and ***Z*** is the embedding in the TP space.

### Model training

The raw scRNA-seq read count data ***X*** is fed into iGTP, which is a matrix of *C* rows (cells) and *Nge_n_e* columns (genes). The training of iGTP follows the training of traditional VAEs by forming a data compression and reconstruction task. The Encoder network to maps the ***X*** to embedding vector ***Z***, and the Decoder neural networks reconstruct ***X̃*** to approximate the input. iGTP is trained by optimizing the reconstruction loss as the KL divergence loss as below,

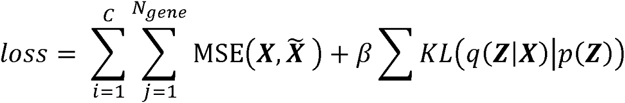

### Measuring Differential TP activity via Plaid-based Bayes Factor

Different treatment conditions often elicit varying activities and TPs. To identify these differential activity genes and/or TPs, we utilize the Bayesian differential gene expression method inspired by Lopez et al.^7^.

The method assumes cell independence and utilizes a Monte Carlo sampling approach by randomly sampling cells from the two treatment groups, denoted as *g_a_,g_b_* respectively. For each TP *q,* the method constructs two hypotheses for any pair of cells *x_a_* ∈ *g_a_* and *x_b_* ∈ *g_b_* to test if the TP *q* has different activations in the two cells from different conditions.

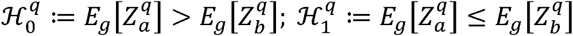

The log-Bayes factor *K* is defined as

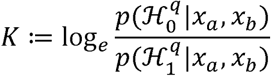

where the sign and magnitude of *K* denote likelihood of each hypothesis and the significance level, respectively.

To achieve robust results, the method aggregates the Bayes factors of a group of cells and computes their average. In iGTP, we use a threshold of 5 *(K* larger than 5) to denote significant differentiation of TP activity. The process of randomly sampling cells from different groups can introduce bias, especially when dealing with the presence of various cell types, some of which may have very small numbers. To address this challenge, we draw inspiration from the Geometric Sketching method proposed by Hie et al.^41^ Geometric sketching is a sampling method to summarize the single-cell transcriptomic landscape. The method highlights rare cell states compared to traditional uniform sampling methods. This approach allows us to subsample large single-cell RNA sequencing (scRNA-seq) datasets while preserving the representation of rare cell states. Geometric Sketching samples cells from cell types according to each cell type’s area of geometric plaid covering in the latent space. The sampling approach is less prone to bias by more evenly sampling according to cell types of landscape, rather than uniform sampling which may underrepresent the rare cell types.

### iGTP perturbation prediction function iGEARS and perturbation embedding integration

We further integrated our TP embedding with a perturbation relationship graph adapted from GEARS^16^ and named iGEARS, to predict the *in silico* impact of gene perturbation on TPs and genes. iGEARS leverages prior knowledge of gene-gene relationships using a GO-derived knowledge graph from GEARS. Intuitively, genes that are involved in similar pathways should impact the expression of similar genes after perturbation. iGEARS functionalizes this graph-based inductive bias using a GNN architecture. Specifically, for cells from perturbation dataset such as Norman et al.^32^, *D =* {*g*_1_, …, *g_NGene_*}, *P*(*c*) | c = 1, …, *C*} where (g_1_,…, *g_NGene_*)*_c_* is the expression of cell *c* and P(c) stands for the perturbation of cell c,from perturbation types ***P*** *=* {*P_1_, …,P_M_*}. We initially applied the unperturbed cells into iGTP Encoder *fi_GTP_·*R*^NGene^ →* R*^NTP^* that maps each cell into TPs of unperturbed cell in *N_TP_*-dimensional embedding. Then, a GNN-based Encoder *f (pert_graph_):**Z** →* R*^d^* is adopted to map each perturbation *P* to a *d-*dimensional perturbation embedding (emb_p_). Lastly, we concatenated the emb_p_ with each TP embedding of unperturbed cells, resulting in an integrated (*d*+1)-dimensional embedding. This integrated embedding was used as input for the cross-TP MLP Decoder *f(_P_ert_dec_):R^*N_Tp_*^* → R*^N_Gene_^* to predict gene expression alteration to unperturbed cells in response to unseen perturbation type *P* (Fig. 1 step 4). We evaluated the prediction performance using top the 20 mean gene expression alteration in perturbated cell P(c) (Fig. 1 step 5).

### Latent diffusion model for conditionally generation of cell types and phenotypes using iGTP and TP embeddings

To further evaluate the generative capabilities of our iGTP model, we integrated its TP embeddings into the latent diffusion model, scDiffusion^37^, which is a generative framework consisting of autoencoder, diffusion process, denoising network, and a conditional classifier. This integration enables the generation of new cells corresponding to specific cell types and phenotypes. Specifically, the pre-trained autoencoder was adapted from iGTP Ahern et al.^38^

PBMC dataset and an single-cell dataset Kang et al. ^17^ was processed through this autoencoder to generate TP embeddings, which were subsequently L2-normalized as latent representations. The diffusion process further produces a series of progressively noisy embeddings, which serve as the training data for the denoising network. In scDiffusion, the denoising network is trained to model the reverse diffusion process, while the classifier is designed to guide the generation toward specific cell types and states. Notably, the classifier is trained on intermediate states derived from the diffusion process, with a particular focus on states at step 0 and step T/2, ensuring robust and accurate conditional generation.

Our experiment consisted of two primary components: training the denoising network and the classifier model. Both models were adapted from the foundational architectures of scDiffusion, with modifications to accommodate the updated embedding and layer dimensions. The denoising network, implemented as a skip-connected multilayer perceptron with an input dimension of 1303 (GO TPs) and hidden layer dimensions of [4096, 2048, 2048], was trained to model the reversed diffusion process. This network was trained for 800,000 steps with a minibatch size of 12 and a learning rate of 1^-4^. The classifier, a feed-forward neural network with an input dimension of 1303 and appropriately sized hidden layers, was trained on noised samples at steps 0 and T/2. Training was conducted with a minibatch size of 256, a learning rate of 5^-5^, and a total of 10,000 steps. After training the scDiffusion model, we generated hundreds of synthetic samples corresponding to specific selected cell types. For additional details, please refer to the scDiffusion^37^.

### Datasets and preprocessing

#### Kang et al. PBMC dataset

The Kang et al. dataset^17^, (accessed by 10/23/2023 from GSE96583), comprises two states of Peripheral Blood Mononuclear Cells (PBMC): a control state and another stimulated with interferon-β. Data was processed by Seurat^42^ and filtered with the following criteria “percent.mt <= 50 & nCount_RNA > 677 & nFeature_RNA > 500 & multiplets == ‘singlet’”. In total, we obtained a gene matrix of 13,672 cells by 35,635 genes. Cell type annotation and states were directly adapted from the original MetaData file in the GSE96583 repository. Megakaryocytes were excluded for their small population. We then converted the Seurat object to Scanpy AnnData object^43^ for further analysis.

#### Ahern et al. COVID-19 PBMC dataset

The Ahern et al.^38^ dataset was downloaded from the Chan Zuckerberg (CZ) CellxGene portal^44^ (accessed by 11/10/2023). The dataset provided an extensive scRNA-seq analysis to construct a blood atlas of patients across a spectrum of COVID-19 severity levels, comparing it with cases of influenza, sepsis, and healthy volunteers. We conducted the standard quality control process using Scanpy^43^. Specifically, we filtered cells with 200 expressed genes and genes expressed in less than three cells. Then, we kept the cell with the following criteria [n_genes_by_counts < 10000; pct_counts_mt < 20; total_counts < 50000]. Cell type annotation was adapted from the original dataset and cell types were renamed with the Kang et al dataset, leading to 599,044 cells in total, including the following sample sources: COVID-19 in-patient critical (COVID_CRIT), COVID-19 in-patient severe (COVID_SEV); COVID-19 in-patient mild (COVID_MILD); COVID_LDN (COVID-19 in-patient with low-dose Naltrexone treatment), Influenza and Normal (healthy volunteer). Detailed description for each phenotype can be found in Ahern et al.^38^.

#### Alzheimer’s Disease single-cell RNA-seq and polygenic risk score process

We adapted snRNA-seq data from Synapse portal (syn2580853, accessed by 4/15/2023), which contains 454 individuals from Religious Orders Study/Memory and Aging Project (ROS/MAP). We obtained the matched whole-genome-sequence (WGS) data from the Synapse portal (syn11724057, accessed by 11/10/2022). LDPred2^45^ was used to estimate the individual’s risk by adapting the variant effect size from Wightman et al. GWAS summary statistics^46^. In total, we obtained 407 individuals with matched snRNA-seq and WGS data. To obtain individuals with similar high-risk PRS backgrounds, we selected the top 20 quantiles of PRS. This subset included 53 high-risk Alzheimer’s disease (AD) cases and 15 high-risk cognitively normal (CN) individuals for comparison. Data was processed by Seurat^42^ and filtered with the following criteria “percent.mt <= 50 & nFeature_RNA > 200”. We adapted the cell type annotation from the original study.

#### Norman et al. CRISPR Perturbation dataset

Norman et al.^32^ raw data was downloaded from scPerturb^47^ processed data (accessed by 11/17/2023), which contained 105 single CRISPR activation (CRISPRa) perturbations with at least 20 cells. We used the same QC process as the *Ahern et al. dataset.* After conducting pretraining on the Norman et al. dataset.

### Transcriptional program curation

We incorporated three TP resources. The GO curation (Biological Process: BP, Molecular Function: MF, and Cellular Component: CC) comprises 1303 non-redundant GO terms from WebGestalt^48,49^ (accessed on 5/7/2021). Lastly, we curated 2082 canonical pathways from three major resources: Kyoto Encyclopedia of Genes and Genomes (KEGG), Reactome, and BioCarta pathways from the Molecular Signatures Database^50^ (MSigDB 7.4 C2 category, accessed on 5/10/2021). The brain and AD-related curation is adapted from our previous work, containing 167 TPs^23^.

### Human protein-protein interaction (PPI) curation

Our human PPI curation included one public database and one in-house curation.

#### Pathway Commons curation

Pathway Commons is an integrated resource of publicly available biological pathways information, biochemical reactions, biomolecular complex assembly, transport, catalysis events, and physical interactions involving proteins, DNA, RNA, and small molecules. We downloaded the Pathway Commons V11 “All.hgnc.txt”, which contains the PPI curations from 18 databases^51^. We extracted the file annotation with human source only and used the annotation of “INTERACTION_TYPE = interaction only”, leading to 485,383 human protein-protein interactions and 17,733 unique proteins.

#### PPI compiled curation

PPI compiled was curated from 6 different resources: BioGRID^48,49,52^, ESCAPE^48^, HINT^53^, IRefIndex^54^, ReactomeFI^55,56^, and STRING^57^. In short, we extracted all the human PPI with physical interaction evidence from each of these resources. Specifically, BioGRID is version “BIOGRID-ALL-3.5.171” was downloaded 4/15/2019. ESCAPE is a database for integrating high content published data collected from human and mouse embryonic stem cells (http://www.maayanlab.net/ESCAPE/). All human interactions were accessed by 4/8/2019. High-quality INTeractomes (HINT) database was curated for PPI with experiment validation and was accessed by 4/6/2019. iRefIndex 15.0 for human “9606.mitab.22012018” was used (accessed by 4/10/2019). ReactomeFI (version 2017) for protein complex was downloaded (accessed by 4/11/2019). The STRING version was downloaded. All the PPIs from six resources were merged, including 419,971 protein-protein interactions and 21,216 unique proteins^55^.

### Benchmark with VEGA and ssGSEA

We followed the procedure of Kang et al. for the PBMC dataset from VEGA^15^. Specifically, we selected the top 7,000 highly variable genes and used GO terms containing at least 10% of these genes to construct the hidden layer. This hidden layer was then used to assess clustering performance and calculate the AUC for ’stimulated’ state label prediction.

We used default setting of “runEscape” function in R package “escape”^58^ to conduct the single sample Gene Set Enrichment Analysis (ssGSEA) analyses^19,20^ for each cell in the single-cell dataset. The resulting cell-by-TP matrix was subsequently used for differential analysis and to calculate the AUC for ’stimulated’ state label prediction.

## DATA AVAILABILITY

All the data generated or analyzed in this study is available from the authors upon reasonable request. The overall framework can be downloaded from https://github.com/davidroad/iGTP.

## ACKNOWLEDGEMENTS

We would like to thank Brisa S. Fernandes from UTHealth at Houston for constructive discussion. The authors acknowledge the High-Performance Computing for Research facility at the Center for Secure Artificial Intelligence for Healthcare, McWilliams School of Biomedical Informatics, University of Texas Health Science Center at Houston, and MD Anderson Cancer Center for providing the computational resources that supported the research presented in this paper.

## FUNDING

This research was partially supported by National Institutes of Health grants awarded to Y.D., L.Y., and Z.Z. (R21AG087299), and to Z.Z (U01AG079847, R03AG077191, R01LM012806, R01DE030122, and R01DE029818). K.H was fully supported by Mulva Family Fund (CFS: 600655-80-120663-19). P.P was partially supported by National Cancer Institutes grant (NCT04947254, NCT03748641). We thanked the resource support from the Cancer Prevention and Research Institute of Texas (CPRIT RP240610). I.K. is a CPRIT summer intern in the Biomedical Informatics, Genomics, and Translational Cancer Research Training Program (BIG-TCR) funded by the Cancer Prevention & Research Institute of Texas (CPRIT RP210045).

## Notes

### Competing Interest Statement

The authors have declared no competing interest.

### Funding Statement

This study was funded by U01AG079847

### Author Declarations

GSE96583, https://cellxgene.cziscience.com/collections/8f126edf-5405-4731-8374-b5ce11f53e82;syn2580853;syn11724057;https://www.nature.com/articles/s41592-023-02144-y

### Summary of Updates

The latest version before submission

